# Disease-Targeted Therapies and eGFR Slope over 2 Years in IgA Nephropathy: A Systematic Review and Network Meta-Analysis

**DOI:** 10.64898/2026.10.29.26366792

**Authors:** Yoshiyasu Tongu, Yuzo Morita, Nozomi Omizo, Daisuke Katagiri

## Abstract

**Background:** Several disease-targeted therapies for IgA nephropathy have reported effects on eGFR slope over 2 years or longer in phase 3 trials, but they have not been compared directly. We compared their effects on the annualized total eGFR slope.

**Methods:** We searched PubMed, Embase, the Cochrane Central Register of Controlled Trials, and ClinicalTrials.gov from January 2018 and reviewed nephrology congress abstracts. Randomized trials reporting the annualized eGFR slope over at least 96 weeks were eligible. Placebo-controlled trials were combined in a frequentist network meta-analysis. The primary outcome was the between-group difference in total eGFR slope; secondary outcomes included the relative reduction in eGFR slope, chronic slope, proteinuria, and safety.

**Results:** Of 1523 records screened, six placebo-controlled trials (2437 participants) were included in the main analysis. Compared with placebo, the difference in total eGFR slope was 5.1 (95% confidence interval [CI], 3.5 to 6.7) ml/min per 1.73 m2 per year for telitacicept, 5.0 (3.6 to 6.5) for atacicept, 4.7 (3.8 to 5.6) for sibeprenlimab, 3.0 (2.0 to 4.0) for iptacopan, 1.8 (0.5 to 3.1) for targeted-release budesonide, and 1.4 (0.5 to 2.3) for atrasentan. The pooled effect of the three agents targeting a proliferation-inducing ligand (APRIL) or B-cell activating factor (BAFF) and APRIL was 4.8 (4.2 to 5.5), and their pooled reduction relative to the control-arm decline was 98% (82% to 113%) versus 34% to 49% for the other agents. Serious adverse events were less frequent with telitacicept and atacicept than with placebo, serious infections were more frequent with iptacopan, and discontinuations were more frequent with targeted-release budesonide.

**Conclusions:** In indirect comparisons, agents targeting APRIL or BAFF/APRIL may provide greater improvements in total eGFR slope over approximately 2 years than the other disease-targeted therapies. One of the three B-cell-directed trials was available only as a conference presentation, and direct comparisons are lacking.

**Key Points:**

- B-cell-directed agents slowed eGFR decline over 2 years by 1.8 to 3.4 ml/min per 1.73 m2 per year more than other targeted agents.
- The B-cell-directed agents reduced the control-arm rate of eGFR decline by 98%, versus 34% to 49% with iptacopan, budesonide, or atrasentan.
- Safety signals differed between agents and observation periods were not uniform across trials, so safety comparisons remain tentative.

## Introduction

IgA nephropathy is the most common primary glomerular disease worldwide, and many patients progress to kidney failure despite renin-angiotensin system blockade and, more recently, sodium-glucose cotransporter 2 (SGLT2) inhibitors [1,2]. Because trials with kidney failure as the primary end point require long follow-up, regulatory agencies have accepted proteinuria reduction at about 9 months as a basis for accelerated approval and the eGFR slope over 2 years as a confirmatory end point [3,4], and most phase 3 trials of disease-targeted therapies for IgA nephropathy have now reported results over 2 years or longer.

These trials have tested agents with different mechanisms of action: targeted-release budesonide, the dual endothelin angiotensin receptor antagonist sparsentan, the endothelin receptor antagonist atrasentan, the complement factor B inhibitor iptacopan, and three agents directed at B cells, namely sibeprenlimab, which inhibits a proliferation-inducing ligand (APRIL), and atacicept and telitacicept, which inhibit both B-cell activating factor (BAFF) and APRIL [5–11]. No head-to-head trials are available. A previous meta-analysis pooled the therapies by drug class [12], but at that time the estimate for B-cell modulating agents rested on the 12-month phase 2 trial of sibeprenlimab and no 2-year data were available for complement inhibitors. Since then, 2-year results have been published for sibeprenlimab, iptacopan, and atacicept, and those of telitacicept were presented at ASN Kidney Week 2026 [8–10,13].

We therefore conducted a systematic review and network meta-analysis of randomized controlled trials to compare the effects of disease-targeted therapies on the annualized total eGFR slope over at least 96 weeks in adults with IgA nephropathy. Because the rate of eGFR decline in the control arm differed between trials and acute effects on eGFR differ by mechanism, we also examined the relative reduction in eGFR slope and the chronic slope, and explored whether differences between agents could be explained by their effects on proteinuria.

## Methods

### Protocol and registration

This systematic review was conducted and reported according to the PRISMA 2020 statement and its extension for network meta-analyses [14,15]. The protocol was registered in PROSPERO (CRD420261548570) before formal screening, data extraction, and risk of bias assessment.

### Data sources and study selection

We included randomized controlled trials in adults with biopsy-proven primary IgA nephropathy and persistent proteinuria despite maximally tolerated renin-angiotensin system blockade, with or without SGLT2 inhibitors, that compared a disease-targeted agent (APRIL inhibitors, dual BAFF/APRIL inhibitors, complement inhibitors, endothelin receptor antagonists, dual endothelin angiotensin receptor antagonists, or targeted-release budesonide) added to supportive care, had a randomized comparison period of at least 96 weeks and reported the annualized eGFR slope with a measure of uncertainty. Trials of systemic immunosuppression or of supportive therapies alone were excluded. Peer-reviewed publications, conference abstracts and posters, and results posted in trial registries were eligible.

We searched PubMed, Embase, and the Cochrane Central Register of Controlled Trials from January 1, 2018, to October 24, 2026, without language restrictions, and ClinicalTrials.gov for phase 2/3 and phase 3 trials in IgA nephropathy started from 2018 (**Supplemental Methods 1**). We also reviewed abstracts from ASN Kidney Week and the ERA Congress from 2024 to 2026 and the reference lists of included trials and relevant reviews, and sought unclear data from investigators where possible.

### Data collection and quality assessment

After deduplication, two reviewers (Y.M. and N.O.) independently screened titles and abstracts and then full texts in Rayyan, with disagreements resolved by discussion or by a third reviewer (D.K.) (**Supplemental Methods 2**). Two reviewers independently extracted trial design, eligibility criteria, baseline characteristics, the definition and statistical model of the eGFR slope, the handling of intercurrent events, efficacy outcomes, and adverse events. When a trial was reported in several sources, we used the most recent report with the longest randomized follow-up. Standard errors were derived from 95% confidence intervals (CIs) when not reported directly.

Two reviewers independently assessed the risk of bias of the total eGFR slope in each trial with the revised Cochrane risk of bias tool (RoB 2) [16], and the certainty of each network estimate was rated with the Confidence in Network Meta-Analysis (CINeMA) framework [17]. Outcomes registered in trial registries and protocols were compared with the reported results.

### Outcomes

The primary outcome was the annualized total eGFR slope over the longest reported double-blind period of at least 96 weeks, estimated from randomization, expressed as the mean difference versus placebo. When several estimates were reported, that from a random-coefficients (mixed-effects) model of all eGFR values from randomization was preferred, and alternative estimates were used in sensitivity analyses. Secondary outcomes were the relative reduction in eGFR slope versus the control arm, the chronic (post-acute) eGFR slope and the acute eGFR effect as defined by each trial, the change in eGFR from baseline to 2 years, the composite kidney outcome of each trial, urine protein-to-creatinine ratio at the primary proteinuria time point, serious adverse events, serious infections, and adverse events leading to discontinuation of the study drug.

### Data synthesis and statistical analyses

A frequentist network meta-analysis was performed with placebo as the common comparator; the main analysis included placebo-controlled trials only. With one trial per comparison, between-study heterogeneity could not be estimated, and a common-effect model was used. Because the network was star-shaped, all comparisons between active agents relied on indirect evidence, and inconsistency could not be assessed. Treatments were ranked with exploratory P-scores [18]. The relative reduction in eGFR slope was calculated as the between-group difference in slope divided by the absolute slope in the control arm, with its standard error derived by the delta method (**Supplemental Methods 3**). The APRIL and BAFF/APRIL inhibitors were pooled as one class of B-cell-directed agents, between-agent heterogeneity was quantified with I2, and the pooled class effect was compared indirectly with each of the other agents.

The plausibility of transitivity was assessed qualitatively by comparing potential effect modifiers across trials, including region, baseline eGFR and proteinuria, SGLT2 inhibitor use, background supportive care, treatment duration, length of the slope window, and handling of intercurrent events; the total slope, chronic slope, and acute eGFR effect were tabulated for each trial where available. Prespecified sensitivity analyses used the chronic eGFR slope where reported, added the active-comparator trial with maximally dosed irbesartan treated as equivalent to placebo plus optimized supportive care, excluded the single-country trial and trials available only as conference presentations, restricted the slope window to about 2 years, used alternative slope estimates, and added a trial whose results were available only from a trial registry (**Supplemental Methods 3**).

Two prespecified exploratory analyses examined the association between proteinuria reduction and the difference in total eGFR slope by trial-level inverse-variance meta-regression, with and without an indicator for B-cell-directed agents, and effect modification by baseline SGLT2 inhibitor use by pooling the within-trial difference between subgroup estimates across trials. Safety outcomes were pooled as risk ratios. Three post hoc analyses were added after registration: network meta-analyses of the change in eGFR at the end of the slope window and of the hazard ratio for each trial’s composite kidney outcome, the latter interpreted only for direction and consistency, and pooling of the within-trial difference in treatment effect between younger and older participants (cut-off 40 or 45 years). Analyses were performed in R version 4.3.3 with the netmeta (version 3.7.0), meta, and metafor packages [19].

## Results

### Study selection and characteristics

The search identified 2321 records, of which 1523 were screened after removal of duplicates and 12 were reviewed in full text (**Supplemental Figure 1**). Six placebo-controlled trials (VISIONARY, APPLAUSE-IgAN, ORIGIN 3, TELIGAN, NefIgArd, and ALIGN) were included in the main analysis [5,7–11,13]; PROTECT (sparsentan versus irbesartan) [6] and the terminated narsoplimab trial ARTEMIS-IGAN, whose 96-week result was available only from ClinicalTrials.gov [20], were included in sensitivity analyses. Data for TELIGAN were obtained from a poster presented at ASN Kidney Week 2026 (TH-PO1256) and data for the other trials from peer-reviewed publications.

The six trials in the main analysis randomized 2437 participants (**Table 1**). TELIGAN was conducted in China; the other trials were multinational. Mean or median baseline eGFR ranged from 55 to 77 ml/min per 1.73 m2, and baseline SGLT2 inhibitor use ranged from 0% in the main stratum of ALIGN to 63% in ORIGIN 3. The slope window was 104 weeks or 24 months in four trials, 2 years in NefIgArd, in which treatment was given for 9 months, and 136 weeks in ALIGN; the slope model and the handling of intercurrent events differed between trials (**Supplemental Table 1**). The risk of bias was low in four trials and in PROTECT, raised some concerns in ALIGN (differential censoring after intercurrent events without a sensitivity analysis), and was high in TELIGAN, in which eGFR values after rescue therapy, started far more often with placebo, were set to missing without a sensitivity analysis (**Supplemental Figure 3** and **Supplemental Table 3**).

**Table 1.** Characteristics of the included trials.

| **Trial** | **Agent (mechanism)** | **Comparator** | **Participants (agent/control)** | **Region** | **Baseline eGFR** | **SGLT2 inhibitor at baseline** | **Slope window** | **Data source** |
|---|---|---|---|---|---|---|---|---|
| TELIGAN | Telitacicept (BAFF/APRIL) | Placebo | 159/159 | China | 77 / 74 (mean) | 35% / 35% | 104 weeks | Poster TH-PO1256, ASN Kidney Week 2026 |
| ORIGIN 3 | Atacicept (BAFF/APRIL) | Placebo | 214/214 | 31 countries | 65 / 63 (mean) | 63% / 60% | 104 weeks | Barratt 2026, NEJM |
| VISIONARY | Sibeprenlimab (APRIL) | Placebo | 259/251 | 29 countries or regions | 65 / 63 (mean) | 43% / 47% | 24 months | Rizk 2026, NEJM |
| APPLAUSE-IgAN | Iptacopan (factor B) | Placebo | 238/239 | 34 countries or regions | 63 / 66 (mean) | 21% / 17% | 24 months | Barratt 2026, NEJM |
| NefIgArd | Targeted-release budesonide | Placebo | 182/182 | 20 countries | 56 / 55 (median) | Not reported (started during the trial: 5% / 6%) | 2 years (9-month treatment) | Lafayette 2023, Lancet |
| ALIGN (main stratum) | Atrasentan (ERA) | Placebo | 170/170 | 20 countries (133 sites) | 58 / 60 (mean) | 0% (by design) | 136 weeks | Heerspink 2026, Lancet |
| PROTECT (sensitivity only) | Sparsentan (DEARA) | Irbesartan | 202/202 | 18 countries (134 sites) | 57 / 57 (mean) | Not permitted | 110 weeks | Rovin 2023, Lancet |

eGFR in ml/min per 1.73 m2. APRIL, a proliferation-inducing ligand; BAFF, B-cell activating factor; DEARA, dual endothelin angiotensin receptor antagonist; ERA, endothelin receptor antagonist; SGLT2, sodium-glucose cotransporter 2. Number of countries from the trial reports: ORIGIN 3, 157 sites in 31 countries (interim report [29]); VISIONARY and APPLAUSE-IgAN, counted from the investigator lists in the supplementary appendices (Hong Kong and Taiwan counted as regions); NefIgArd, ALIGN, and PROTECT, as stated in the Lancet reports.

### Total eGFR slope

Compared with placebo, all six agents slowed the decline in eGFR (**Figure 1**). The difference in total eGFR slope was 5.1 (95% CI, 3.5 to 6.7) ml/min per 1.73 m2 per year for telitacicept, 5.0 (3.6 to 6.5) for atacicept, 4.7 (3.8 to 5.6) for sibeprenlimab, 3.0 (2.0 to 4.0) for iptacopan, 1.8 (0.5 to 3.1) for targeted-release budesonide, and 1.4 (0.5 to 2.3) for atrasentan; P-scores were highest for the three B-cell-directed agents (**Supplemental Table 6**). Pooled as a class, these agents slowed eGFR decline by 4.8 (4.2 to 5.5) ml/min per 1.73 m2 per year, with no observed heterogeneity between agents (I2 = 0%), and in indirect comparisons the class effect was larger than that of iptacopan by 1.8 (0.6 to 3.0), targeted-release budesonide by 3.0 (1.5 to 4.5), and atrasentan by 3.4 (2.3 to 4.6) ml/min per 1.73 m2 per year. Excluding TELIGAN (high risk of bias) did not change the other estimates (**Supplemental Table 2**). Certainty of evidence was high to low versus placebo (low for telitacicept) and very low to moderate between agents (**Supplemental Table 7**).

**Figure 1.**
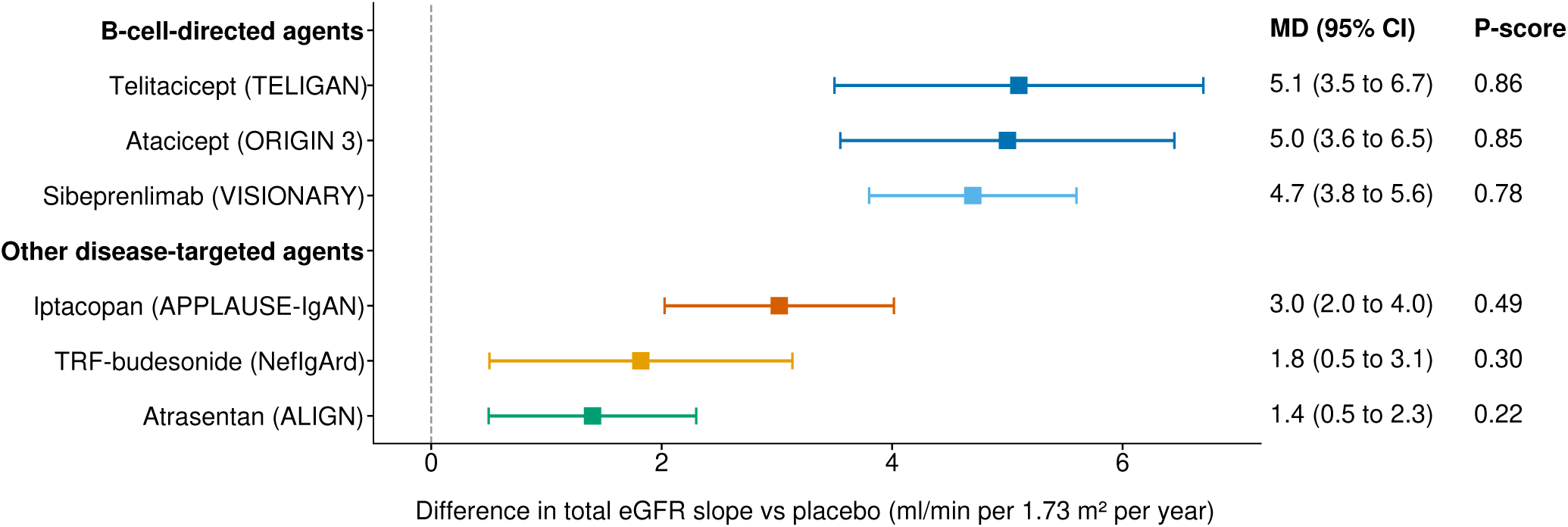
Network meta-analysis of the difference in annualized total eGFR slope versus placebo (common-effect model). Squares indicate point estimates and horizontal lines 95% CIs; positive values indicate slower eGFR decline than with placebo. Agents are grouped by mechanism of action, and P-scores are shown on the right. CI, confidence interval; MD, mean difference; TRF-budesonide, targeted-release budesonide.

### Relative reduction in eGFR slope

The annualized eGFR slope in the control arm ranged from −4.1 to −6.1 ml/min per 1.73 m2 per year (**Figure 2**). Relative to the control arm, the rate of eGFR decline was reduced by 89% (95% CI, 72% to 107%) with telitacicept, 89% (71% to 108%) with atacicept, 112% (97% to 127%) with sibeprenlimab, 49% (37% to 62%) with iptacopan, 34% (13% to 55%) with targeted-release budesonide, and 34% (16% to 52%) with atrasentan; a reduction above 100% indicates a slight increase in eGFR in the treatment arm (0.5 ml/min per 1.73 m2 per year with sibeprenlimab). The pooled relative reduction for the B-cell-directed agents was 98% (82% to 113%), 48 (28 to 68) percentage points larger than with iptacopan and 64 points larger than with targeted-release budesonide or atrasentan.

**Figure 2.**
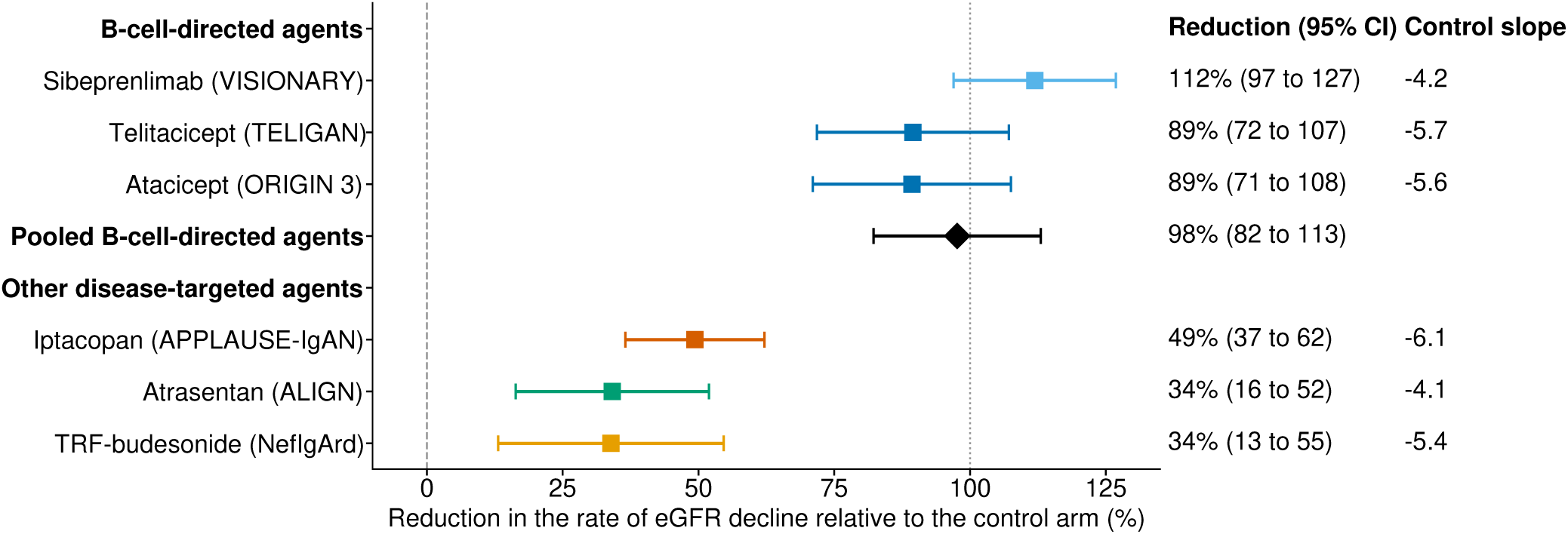
Relative reduction in the annualized eGFR slope versus the control arm in each trial, with the pooled estimate for B-cell-directed agents (random-effects model, diamond). The reduction is the between-group difference in slope divided by the absolute slope in the control arm; the dotted line at 100% corresponds to no eGFR decline in the treatment arm. Squares indicate point estimates and horizontal lines 95% CIs. CI, confidence interval; Control slope, annualized eGFR slope in the control arm (ml/min per 1.73 m2 per year); TRF-budesonide, targeted-release budesonide.

### Chronic slope and acute eGFR effects

Chronic eGFR slope was reported in VISIONARY, ALIGN, and PROTECT (**Table 2**). The difference in chronic slope was 4.3 (3.3 to 5.3) ml/min per 1.73 m2 per year for sibeprenlimab, slightly smaller than the total slope, 1.7 (0.7 to 2.8) for atrasentan, slightly larger than the total slope with no acute reduction in eGFR reported, and 1.1 (0.1 to 2.1) for sparsentan, with a difference in the change in eGFR to week 6 of 0.4 (−1.0 to 1.7). Using the chronic slope where reported did not change the ranking of agents.

**Table 2.** Total slope, chronic slope, and acute eGFR effects.

| **Trial** | **Agent** | **Total slope difference (95% CI)** | **Chronic slope difference (95% CI)** | **Definition of chronic slope** | **Acute eGFR effect** |
|---|---|---|---|---|---|
| VISIONARY | Sibeprenlimab | 4.7 (3.8 to 5.6) | 4.3 (3.3 to 5.3) | Linear spline, knot at week 12 | Slope from week 4: 4.5 (3.6 to 5.4) |
| ALIGN | Atrasentan | 1.4 (0.5 to 2.3) | 1.7 (0.7 to 2.8) | Two-piece slope, knot at week 6 | No acute reduction reported by the investigators |
| PROTECT | Sparsentan | 1.0 (0.0 to 1.9) | 1.1 (0.1 to 2.1) | Weeks 6 to 110 | Change to week 6: 0.4 (-1.0 to 1.7) |
| APPLAUSE-IgAN | Iptacopan | 3.0 (2.0 to 4.0) | Not reported |  | Not reported |
| NefIgArd | Targeted-release budesonide | 1.8 (0.5 to 3.1) | Not reported |  | Not reported |
| ORIGIN 3 | Atacicept | 5.0 (3.6 to 6.5) | Not reported |  | Not reported |
| TELIGAN | Telitacicept | 5.1 (3.5 to 6.7) | Not reported |  | Not reported |

Values in ml/min per 1.73 m2 per year (difference versus control). CI, confidence interval.

### Sensitivity analyses

Restriction to slope windows of about 2 years, use of the post hoc intention-to-treat slope of ORIGIN 3 (atacicept 4.2; 2.8 to 5.5), and derivation of the NefIgArd slope from the change in eGFR at 2 years (targeted-release budesonide 3.0; 1.5 to 4.4) did not change the direction or ranking of the estimates (**Supplemental Table 2**). Adding the active-comparator trial gave a difference of 1.0 (0.0 to 1.9) ml/min per 1.73 m2 per year for sparsentan without changing the other estimates, and adding ARTEMIS-IGAN gave 1.2 (−0.7 to 3.1) for narsoplimab.

### Change in eGFR and composite kidney outcomes

The change in eGFR at the end of the slope window, reported for four trials, differed from placebo by 9.2 (7.1 to 11.4) ml/min per 1.73 m2 for sibeprenlimab, 5.0 (2.8 to 7.2) for iptacopan, 5.9 (3.2 to 8.6) for targeted-release budesonide, and 2.4 (−0.1 to 4.9) for atrasentan (**Supplemental Figure 6**). All six trials reported a composite kidney outcome, with definitions that differed between trials (**Supplemental Table 4**); the hazard ratios versus placebo ranged from 0.03 (0.004 to 0.21) for sibeprenlimab to 0.64 (0.39 to 1.05) for atrasentan (**Supplemental Figure 7**). The pooled hazard ratio for the B-cell-directed agents was 0.13 (0.05 to 0.34; I2 = 57%), lower in indirect comparisons than that of iptacopan (ratio of hazard ratios, 0.22; 0.08 to 0.63), atrasentan (0.20; 0.06 to 0.59), and targeted-release budesonide (0.28; 0.09 to 0.86). In PROTECT, the composite kidney failure end point occurred in 9% versus 13% (relative risk, 0.7; 0.4 to 1.2).

### Proteinuria and eGFR slope

Across seven trials including PROTECT, each 10% greater reduction in urine protein-to-creatinine ratio was associated with a 1.7 (1.1 to 2.3) ml/min per 1.73 m2 per year larger difference in total eGFR slope (**Figure 3**). After adding an indicator for B-cell-directed agents, the association with proteinuria reduction was attenuated (−0.2; −1.2 to 0.9), and the B-cell-directed agents were associated with a 3.3 (1.8 to 4.8) ml/min per 1.73 m2 per year larger effect on eGFR slope after adjustment for proteinuria reduction. Given the small number of trials and the potential for ecological bias, this analysis is hypothesis-generating.

**Figure 3.**
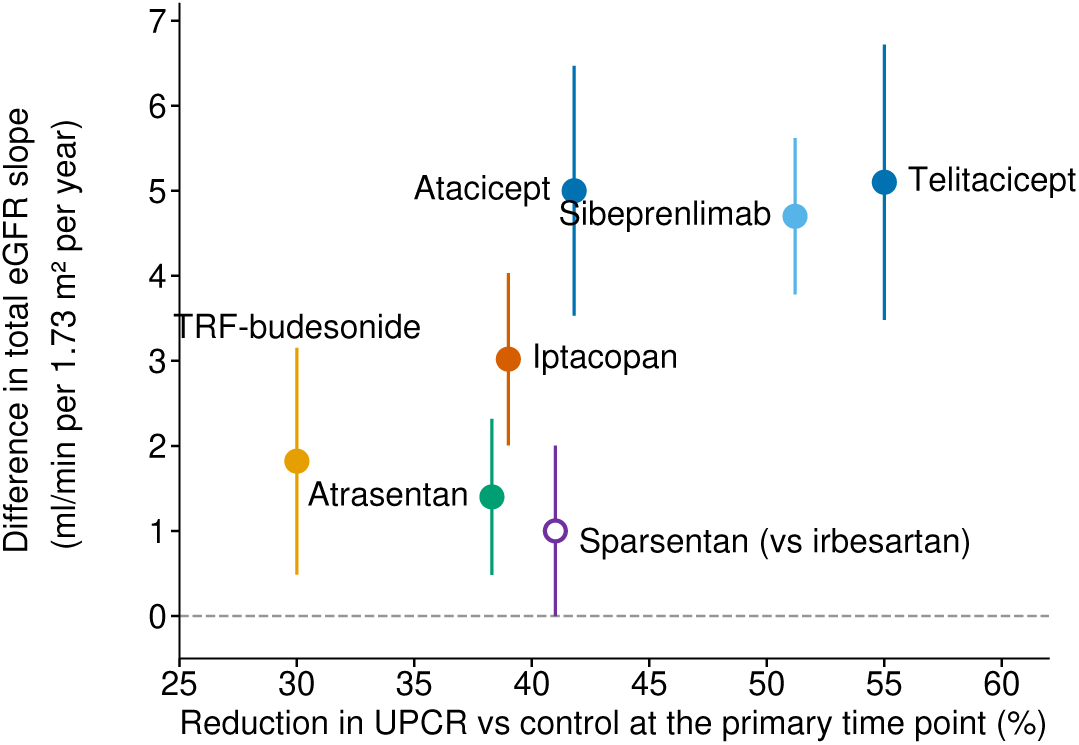
Trial-level association between the reduction in urine protein-to-creatinine ratio at the primary proteinuria time point and the difference in total eGFR slope. Each circle represents one trial, and vertical lines indicate 95% CIs of the difference in total eGFR slope; the open circle indicates the active-comparator trial (sparsentan versus irbesartan). CI, confidence interval; TRF-budesonide, targeted-release budesonide; UPCR, urine protein-to-creatinine ratio.

### Subgroups

Subgroup estimates by baseline SGLT2 inhibitor use were available for ORIGIN 3, VISIONARY, APPLAUSE-IgAN, and ALIGN (**Figure 4**); the difference in treatment effect between participants with and without SGLT2 inhibitors was 0.9 (−0.2 to 2.1) ml/min per 1.73 m2 per year, with no heterogeneity (I2 = 0%). In the same trials, the pooled differences in treatment effect by baseline proteinuria (higher versus lower), baseline eGFR, region, and sex were 1.0 (−0.1 to 2.0), −0.6 (−1.6 to 0.4), 0.6 (−0.4 to 1.6), and −0.2 (−1.2 to 0.8) ml/min per 1.73 m2 per year (**Supplemental Figure 4** and **Supplemental Table 5**). By age (cut-off 40 or 45 years), the effect was larger in younger participants for atacicept (6.6 versus 3.7) and sibeprenlimab (5.6 versus 3.7) but not for iptacopan (2.7 versus 3.7); the pooled difference was 0.9 (−0.3 to 2.2) across the three trials (I2 = 70%) and 2.2 (0.6 to 3.8) for the two B-cell-directed agents (**Supplemental Figure 5**).

**Figure 4.**
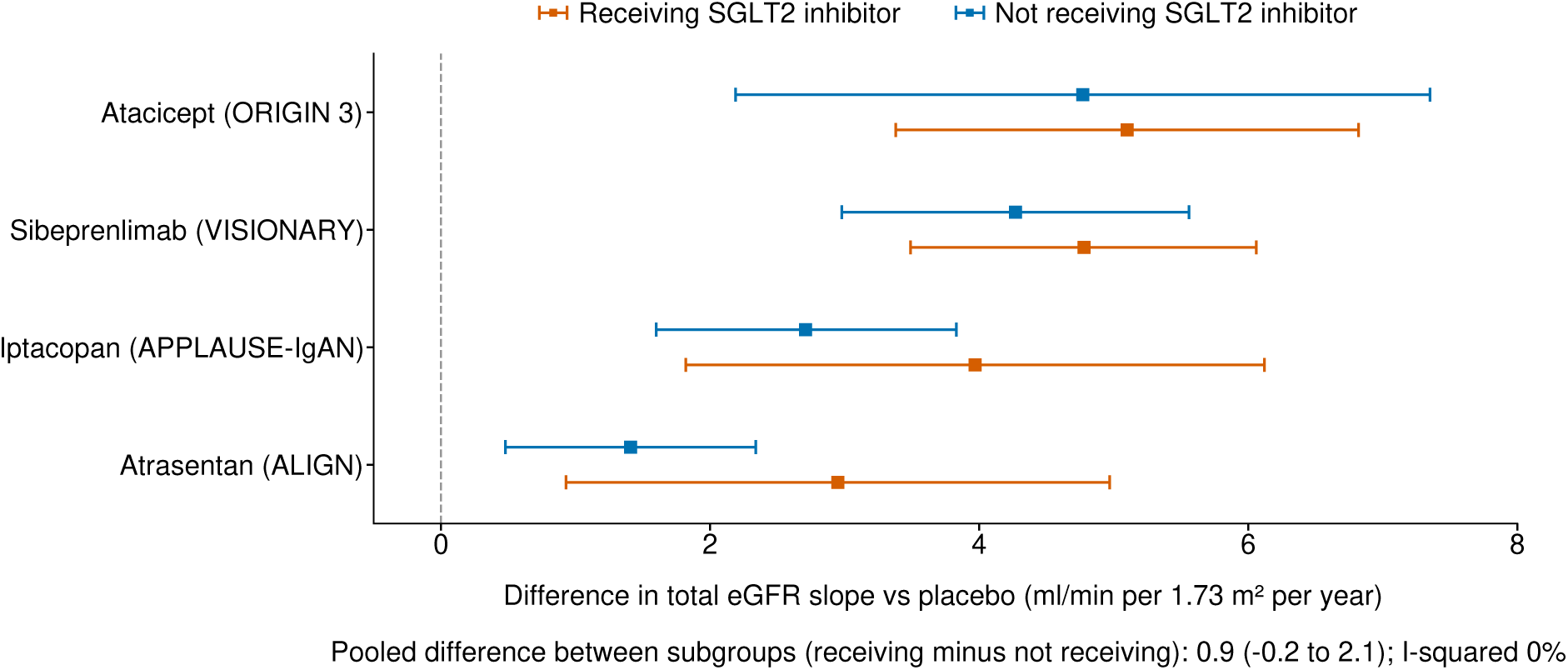
Difference in total eGFR slope by baseline SGLT2 inhibitor use in ORIGIN 3, VISIONARY, APPLAUSE-IgAN, and ALIGN. Squares indicate point estimates and horizontal lines 95% CIs in participants receiving (orange) and not receiving (blue) an SGLT2 inhibitor at baseline; for ALIGN, the SGLT2 inhibitor stratum is shown against the main stratum. The pooled within-trial difference between subgroups is shown below the plot. CI, confidence interval; SGLT2, sodium-glucose cotransporter 2.

### Safety

Compared with placebo, serious adverse events were less frequent with telitacicept (risk ratio, 0.45; 95% CI, 0.24 to 0.83) and atacicept (0.45; 0.25 to 0.81) and did not differ significantly for the other agents (**Table 3**). Serious infections were more frequent with iptacopan (3.21; 1.20 to 8.63), with wide CIs including 1 for the other agents. Adverse events leading to discontinuation were less frequent with atacicept (0.18; 0.06 to 0.52) and more frequent with targeted-release budesonide (5.67; 1.69 to 19.0). In PROTECT, serious adverse events (37% versus 35%) and discontinuations because of adverse events (10% versus 9%) were similar with sparsentan and irbesartan.

**Table 3.** Safety outcomes versus placebo (risk ratio, 95% CI)

| **Agent** | **Serious adverse events** | **Serious infections** | **Adverse events leading to discontinuation** |
|---|---|---|---|
| Telitacicept | 0.45 (0.24 to 0.83) | 0.43 (0.11 to 1.63) | 0.43 (0.11 to 1.63) |
| Atacicept | 0.45 (0.25 to 0.81) | 0.75 (0.26 to 2.12) | 0.18 (0.06 to 0.52) |
| Sibeprenlimab | 0.64 (0.38 to 1.07) | 0.48 (0.17 to 1.40) | 0.29 (0.08 to 1.04) |
| Iptacopan | 1.04 (0.64 to 1.69) | 3.21 (1.20 to 8.63) | 1.00 (0.44 to 2.27) |
| Targeted-release budesonide | 2.00 (0.92 to 4.33) | 2.50 (0.49 to 12.72) | 5.67 (1.69 to 19.00) |
| Atrasentan | 0.93 (0.56 to 1.55) | Not reported | 0.60 (0.22 to 1.62) |

Risk ratios versus placebo from common-effect network meta-analysis of the six placebo-controlled trials. Events were counted over the double-blind period of each trial (approximately 2 years), except for NefIgArd, where serious adverse events and discontinuations were reported for the 9-month treatment period only. Serious infections in ORIGIN 3 were reported as serious or severe infections, and in NefIgArd as serious infection-related adverse events. PROTECT (sparsentan versus irbesartan) was not pooled. CI, confidence interval.

## Discussion

In this systematic review and network meta-analysis of six placebo-controlled phase 3 trials, all disease-targeted therapies slowed the decline in eGFR over 2 years or longer, but the size of the effect differed by mechanism of action. The three agents directed at B cells reduced the total eGFR slope by 4.7 to 5.1 ml/min per 1.73 m2 per year, with no heterogeneity between agents, whereas the effects of iptacopan, targeted-release budesonide, and atrasentan ranged from 1.4 to 3.0; relative to the placebo arms, the reductions were about 90% or more and 34% to 49%, respectively. Telitacicept and atacicept were also associated with fewer serious adverse events than placebo, whereas serious infections were more frequent with iptacopan and discontinuations with targeted-release budesonide.

Our findings agree with a previous meta-analysis that pooled therapies by drug class and reported a larger effect on eGFR slope for B-cell modulating agents (4.3 ml/min per 1.73 m2 per year) than for corticosteroids (2.3) or nonimmunologic therapies (1.1) [12], although its estimate for B-cell modulating agents rested on the 12-month phase 2 trial of sibeprenlimab. To our knowledge, the present analysis is the first to compare individual agents within a network using the 2-year phase 3 results of all three B-cell-directed agents, including those of telitacicept presented in October 2026. Because the annualized slope in the placebo arms ranged from −4.1 to −6.1 ml/min per 1.73 m2 per year, we also expressed the effects relative to the control arm, as in previous trials and meta-analyses [4,12]; the ranking was the same on both scales.

APRIL and BAFF support the survival of the B cells and plasma cells that produce galactose-deficient IgA1, and inhibition of these cytokines lowers galactose-deficient IgA1 and immune-complex formation [21,22]. The effect of the three B-cell-directed agents was consistent despite differences in molecule, dosing, and trial population, and no increase in serious infections was observed over 2 years. However, these agents lower serum immunoglobulins, and the consequences of long-term treatment, including infections, hypogammaglobulinemia, and the response to vaccination, cannot be assessed from 2-year trials. Galactose-deficient IgA1 returned toward baseline after sibeprenlimab was stopped in the phase 2 trial [22], so the duration of treatment needed to maintain the benefit is unknown.

The effect of iptacopan was intermediate. Alternative pathway activation contributes to glomerular injury downstream of immune-complex deposition, and factor B inhibition does not act on the production of galactose-deficient IgA1 [8,23]. Serious infections were more frequent with iptacopan, in keeping with the known risk of infection with encapsulated bacteria during complement inhibition, so vaccination and surveillance are required [24]. The effects of targeted-release budesonide and atrasentan were smaller. In NefIgArd, treatment was given for 9 months, so the 2-year slope includes 15 months off treatment, and the slope during treatment was steeper than after treatment [5]. Atrasentan and sparsentan act on glomerular hemodynamics, and their effects were similar to each other and to that of SGLT2 inhibitors in the previous meta-analysis (1.1 ml/min per 1.73 m2 per year) [12].

The total eGFR slope includes any acute effect of treatment on eGFR, which differs by mechanism [4,25,26]. Agents with hemodynamic actions may lower eGFR early after initiation, making the total slope smaller than the chronic slope. In the trials that reported both measures, the chronic slope was slightly larger than the total slope for atrasentan and sparsentan and slightly smaller for sibeprenlimab [7], and the ranking of agents was unchanged. However, because the chronic slope was not reported for four of the six trials, differences in acute effects cannot be fully excluded as an explanation for the gap between the B-cell-directed agents and the other agents. In the trial-level meta-regression, the reduction in proteinuria was associated with the effect on eGFR slope across agents, in agreement with patient-level analyses [3,4], but the B-cell-directed agents had larger effects on eGFR slope after adjustment for proteinuria reduction. This pattern may reflect effects of lowering galactose-deficient IgA1 that are not captured by proteinuria, but also differences in trial populations and in the timing of proteinuria measurement. There was no clear evidence that the effects of four agents differed by baseline SGLT2 inhibitor use, proteinuria, eGFR, region, or sex, suggesting that the effects may be maintained with SGLT2 inhibitors, although the subgroup analyses were not powered to show equivalence. One exception was age: the effects of atacicept and sibeprenlimab were about 2 ml/min per 1.73 m2 per year larger in participants younger than 40 years, whereas no such difference was seen with iptacopan. This observation rests on unadjusted subgroup estimates from two trials and should be regarded as hypothesis-generating.

Which agent should be used for which patient cannot be answered from indirect comparisons; patient-level data sharing across trials would allow comparisons within subgroups. The phase 3 trials of povetacicept and ravulizumab have so far reported only interim proteinuria results [27,28]. Whether the agents act additively has not been tested, the optimal duration of treatment is unknown, and longer follow-up is needed to confirm that differences in eGFR slope translate into differences in kidney failure.

The strengths of this study include the prospectively registered protocol, the restriction of the main analysis to placebo-controlled phase 3 trials reporting the eGFR slope over at least 96 weeks, and the prespecified sensitivity analyses. There are limitations. First, the network was star-shaped, so all comparisons between active agents were indirect, and inconsistency could not be assessed. Second, the 2-year results of TELIGAN were available only as a conference poster and were at high risk of bias from differential missing data; excluding TELIGAN did not change the other estimates. Third, the trials differed in population, background therapy, treatment duration, and slope methodology, so comparative effectiveness should be interpreted with caution. Fourth, the eGFR slope is a surrogate end point, and follow-up was too short to compare effects on kidney failure; the composite kidney outcomes pointed in the same direction, but definitions differed, events were few, and the CI for sibeprenlimab was very wide, so this comparison is only supportive. Fifth, safety data, particularly on infections with long-term B-cell-directed therapy, were limited to 2 years, and the safety events of NefIgArd were reported for the 9-month treatment period only.

In conclusion, indirect comparisons of phase 3 trials suggest that agents targeting APRIL or BAFF/APRIL may provide greater improvements in total eGFR slope than the other disease-targeted therapies evaluated for IgA nephropathy. However, differences in trial characteristics and eGFR slope estimands limit the certainty of these comparisons.

## Data Availability

Original data generated for the study are provided in the article and its Supplemental Material. Data Type: Published Material. The analysis code (R) will be made available upon reasonable
request to the corresponding author.

## Acknowledgments

None.

## Author Contributions

Conceptualization: Y. Tongu, D. Katagiri. Methodology: Y. Tongu. Investigation: Y. Morita, N. Omizo, Y. Tongu. Data curation: Y. Morita, N. Omizo. Formal analysis: Y. Tongu. Validation: Y. Morita, N. Omizo. Visualization: Y. Tongu. Writing, original draft: Y. Tongu. Writing, review and editing: Y. Morita, N. Omizo, D. Katagiri. Supervision and project administration: D. Katagiri.

## Data Sharing Statement

Original data generated for the study are provided in the article and its Supplemental Material. Data Type: Published Material. The analysis code (R) will be made available upon reasonable request to the corresponding author.

## Supplemental Material

This article contains the following supplemental material:

Supplemental Methods 1. Search strategies.

Supplemental Methods 2. Screening criteria.

Supplemental Methods 3. Statistical details and sensitivity analyses.

Supplemental Table 1. Design, eGFR slope model and handling of intercurrent events in the included trials.

Supplemental Table 2. Sensitivity analyses of the difference in total eGFR slope versus placebo.

Supplemental Table 3. Risk of bias (RoB 2) for the total eGFR slope.

Supplemental Table 4. Definitions of the composite kidney outcomes and proteinuria remission end points.

Supplemental Table 5. Within-trial subgroup estimates of the difference in total eGFR slope versus placebo.

Supplemental Table 6. League table of the main analysis. Supplemental Table 7. Certainty of evidence (CINeMA).

Supplemental Figure 1. PRISMA 2020 flow diagram of study selection.

Supplemental Figure 2. Network of the main analysis.

Supplemental Figure 3. Risk of bias (RoB 2) for the total eGFR slope in each trial.

Supplemental Figure 4. Difference in total eGFR slope versus placebo by baseline proteinuria, baseline eGFR, region and sex.

Supplemental Figure 5. Difference in total eGFR slope versus placebo by age.

Supplemental Figure 6. Difference in the change in eGFR from baseline at the end of the slope window versus placebo.

Supplemental Figure 7. Hazard ratios versus placebo for the composite kidney outcome of each trial.

