## Supplemental File for "Disease-Targeted Therapies and eGFR Slope over 2 Years in IgA Nephropathy: A Systematic Review and Network Meta-Analysis"

#### Table of contents

Supplemental Methods 1. Search strategies

Supplemental Methods 2. Screening criteria

Supplemental Methods 3. Statistical details and sensitivity analyses

Supplemental Table 1. Design, eGFR slope model and handling of intercurrent events in the included trials

Supplemental Table 2. Sensitivity analyses of the difference in total eGFR slope versus placebo

Supplemental Table 3. Risk of bias (RoB 2) for the total eGFR slope

Supplemental Table 4. Definitions of the composite kidney outcomes and proteinuria remission end points

Supplemental Table 5. Within-trial subgroup estimates of the difference in total eGFR slope versus placebo

Supplemental Table 6. League table of the main analysis

Supplemental Table 7. Certainty of evidence (CINeMA)

Supplemental Figure 1. PRISMA 2020 flow diagram of study selection

Supplemental Figure 2. Network of the main analysis

Supplemental Figure 3. Risk of bias (RoB 2) for the total eGFR slope in each trial

Supplemental Figure 4. Difference in total eGFR slope versus placebo by baseline proteinuria, baseline eGFR, region and sex

Supplemental Figure 5. Difference in total eGFR slope versus placebo by age

Supplemental Figure 6. Difference in the change in eGFR from baseline at the end of the slope window versus placebo

Supplemental Figure 7. Hazard ratios versus placebo for the composite kidney outcome of each trial

#### Supplemental Methods 1. Search strategies

Two concept blocks were combined: (A) IgA nephropathy and (B) randomized trial design. Intervention and outcome terms were not used, because reports whose abstracts do not name the outcome would otherwise be missed. Eligibility for a randomized comparison period of 2 years or longer, agent class and reporting of the eGFR slope was applied at screening. Block A used controlled vocabulary (MeSH, Emtree) and free-text synonyms including the abbreviation IgAN and the eponym Berger disease. Block B used the Cochrane Highly Sensitive Search Strategy for MEDLINE (sensitivity- and precision-maximizing version) and the Cochrane Embase RCT filter; no design filter was applied in CENTRAL. Records were limited to publication years 2018 onwards, without language restriction. All searches were run on October 24, 2026.

**MEDLINE (PubMed), 326 records**

#1 "Glomerulonephritis, IGA"[Mesh] OR "IgA nephropathy"[tiab] OR "IgA nephropathies"[tiab] OR "immunoglobulin A nephropathy"[tiab] OR "IgA glomerulonephritis"[tiab] OR IgAN[tiab] OR "Berger disease"[tiab] OR "Berger's disease"[tiab]

#2 randomized controlled trial[pt] OR controlled clinical trial[pt] OR randomized[tiab] OR randomised[tiab] OR placebo[tiab] OR "clinical trials as topic"[mesh:noexp] OR randomly[tiab] OR trial[ti]

#3 #1 AND #2; #4 #3 NOT (animals[mh] NOT humans[mh]); #5 #4 AND 2018:2026[dp]

Validation: the strategy retrieved all eight known eligible or related trial reports indexed in PubMed at the time of testing (ORIGIN 3 interim, VISIONARY interim, TELIGAN interim, APPLAUSE-IgAN final, ALIGN final, PROTECT interim and 2-year, NefIgArd 2-year). An earlier draft with an outcome block (eGFR or kidney function terms) retrieved 78 records but missed the ORIGIN 3 interim report, whose abstract did not mention kidney function; the outcome block was therefore removed.

**Embase (Ovid; Embase 1974 to 2026 October 20), 1,170 records**

1 exp immunoglobulin A nephropathy/

2 (iga nephropathy or iga nephropathies or immunoglobulin a nephropathy or iga glomerulonephritis or igan or berger disease or berger* disease).ti,ab,kw.

3 1 or 2

4 exp randomized controlled trial/ or controlled clinical trial/ or exp phase 3 clinical trial/ or double blind procedure/ or (random* or placebo*).ti,ab,tt. or double blind.ti,ab. or trial.ti.

5 3 and 4; 6 5 not (exp animal/ not exp human/); 7 limit 6 to yr="2018-Current"

The protocol was written for Embase.com and translated to Ovid syntax ('/exp' to exp .../; ':ti,ab,kw' to .ti,ab,kw.; [animals]/lim NOT [humans]/lim to exp animal/ not exp human/; [2018-2026]/py to limit yr="2018-Current"). Conference abstracts were not excluded. Validation: DOI look-up of the seven known primary reports found six indexed, all retrieved by line 7 (the VISIONARY final report was not yet indexed).

**Cochrane Central Register of Controlled Trials (Cochrane Library), 786 records**

#1 MeSH descriptor: [Glomerulonephritis, IGA] explode all trees

#2 ("IgA nephropathy" OR "IgA nephropathies" OR "immunoglobulin A nephropathy" OR "IgA glomerulonephritis" OR IgAN OR "Berger disease" OR "Berger's disease"):ti,ab,kw

#3 #1 OR #2; #4 #3 with publication date from January 2018, in Trials

**ClinicalTrials.gov, 39 records**

Condition: IgA Nephropathy OR IgA Glomerulonephritis; study type: interventional; phase: phase 2/phase 3 and phase 3; study start from January 1, 2018; all recruitment statuses. The WHO International Clinical Trials Registry Platform was not searched separately because its records are included in CENTRAL.

**Other sources**

Abstract books of ASN Kidney Week and the ERA Congress (2024 to 2026) were searched by hand for phase 3 trials reporting eGFR outcomes over 2 years or longer; reference lists of included trials and relevant systematic reviews were checked; corresponding authors or sponsors were contacted for unpublished or unclear data.

**Deduplication**

Records were merged on the PubMed identifier, then the DOI, then the normalized title and publication year (2,321 records, including 39 from ClinicalTrials.gov; 798 duplicates removed; 1,523 unique records screened).

#### Supplemental Methods 2. Screening criteria

Titles and abstracts were screened in Rayyan by two reviewers (Y.M. and N.O.) blinded to each other's decisions; disagreements were resolved by discussion or by a third reviewer (D.K.). A record was included only if all four criteria were met: (1) primary IgA nephropathy in adults or mixed ages; (2) a phase 3 (or phase 2/3) randomized controlled trial against placebo or an active comparator; (3) a disease-targeted therapy (APRIL or BAFF/APRIL inhibitor, complement inhibitor, endothelin receptor antagonist or dual endothelin angiotensin receptor antagonist, targeted-release budesonide, or another new targeted agent); and (4) eGFR reported over 2 years or longer (96 weeks or more) of randomized comparison, as a slope or a change. Any report of an included trial (full paper, conference abstract or secondary analysis) that gave 2-year eGFR data was also included. Records were excluded with one reason, applied in this order: E1 not IgA nephropathy; E2 not a randomized trial (review, meta-analysis, editorial, letter, case report, cohort, real-world study, pooled post hoc analysis or guideline); E3 animal or in vitro study; E4 not a disease-targeted therapy (renin-angiotensin system inhibitors, SGLT2 inhibitors, mineralocorticoid receptor antagonists, systemic glucocorticoids, mycophenolate, cyclophosphamide, hydroxychloroquine, tonsillectomy, traditional Chinese medicine, fish oil or statins); E5 phase 1 or phase 2 trial; E6 follow-up shorter than 96 weeks or no 2-year eGFR result; E7 interim or short-term report of an included trial (retained for design and risk-of-bias assessment); E8 ongoing trial, protocol or design paper without results; E9 published before 2018. "Maybe" was used only when no abstract was available and the title alone could not decide. The interim reports of TELIGAN and ORIGIN 3 were linked to those trials as secondary reports and were used only for trial characteristics and risk-of-bias assessment, not for effect estimates; they are counted among the reports of included studies in Supplemental Figure 1.

#### Supplemental Methods 3. Statistical details and sensitivity analyses

**Relative reduction in eGFR slope.** The relative reduction was calculated as the between-group difference in annualized total eGFR slope divided by the absolute value of the control-arm slope. Its standard error was derived by the delta method, taking into account the covariance between the between-group difference and the control-arm slope. When the confidence interval of the control-arm slope was not reported, its standard error was approximated as the standard error of the between-group difference divided by the square root of 2.

**Prespecified sensitivity analyses.** (1) Use of the chronic (post-acute) eGFR slope instead of the total slope where reported; (2) addition of the active-comparator trial (PROTECT), with maximally dosed irbesartan treated as equivalent to placebo plus optimized supportive care; (3) exclusion of the single-country trial (TELIGAN); (4) exclusion of trials available only as conference presentations (TELIGAN); (5) restriction to trials with a slope window of approximately 2 years (exclusion of ALIGN, 136 weeks);

(6) use of alternative slope estimates (the post hoc intention-to-treat slope of ORIGIN 3 with a treatment-policy strategy for all intercurrent events, and the total slope of NefIgArd derived from the change in eGFR at 2 years); (7) exclusion of each trial in turn where relevant; and (8) addition of a trial whose results were available only from a trial registry and could not be verified against the prespecified analysis (ARTEMIS-IGAN, narsoplimab; terminated after an interim analysis, with the 96-week annualized eGFR change posted in ClinicalTrials.gov based on last observation carried forward). Results are given in Supplemental Table 2.

#### Supplemental Table 1. Design, eGFR slope model and handling of intercurrent events in the included trials

| **Trial (agent)** | **Design and sites** | **Primary end point** | **Slope end point and window** | **Slope model** | **Intercurrent events and missing data** | **Source of the slope estimate used** |
|---|---|---|---|---|---|---|
| TELIGAN (telitacicept 240 mg weekly) | Phase 3, double-blind, placebo-controlled; 72 sites in China; n = 318 | Annualized total eGFR slope through week 104 (final analysis); UPCR at week 39 (interim) | Total slope, randomization to week 104 | Longitudinal mixed model for repeated measures with treatment, time and interaction (poster); protocol v3.0 planned per-patient linear regression slopes | Data after rescue or prohibited medication not used (hypothetical strategy); data after discontinuation of the study drug retained; no imputation | Poster TH-PO1256, ASN Kidney Week 2026; NEJM 2026 interim report and protocol for design |
| ORIGIN 3 (atacicept 150 mg weekly) | Phase 3, double-blind, placebo-controlled; 157 sites in 31 countries; n = 428 | UPCR at week 36 | Total slope, first injection to week 104 (key secondary) | Random-coefficients model with random intercepts and slopes; fixed effects for strata, treatment, time and treatment-by-time | Treatment-policy strategy for all intercurrent events; jump-to-reference after study discontinuation; worst case after dialysis, transplantation or death; post hoc ITT slope (4.15) also reported | NEJM 2026 final report (Table 2) |
| VISIONARY (sibeprenlimab 400 mg SC monthly) | Phase 3, double-blind, placebo-controlled; 29 countries or regions; n = 510 | UPCR at 9 months | Total slope from baseline to 24 months (key secondary); slope from week 4 also prespecified | Mixed-effects slope model | Hybrid estimand with hypothetical strategy for rescue medication; treatment-policy sensitivity analysis reported and consistent; missing data assumed missing at random | NEJM 2026 final report (slope from baseline) |
| APPLAUSE-IgAN (iptacopan 200 mg twice daily) | Phase 3, double-blind, placebo-controlled; 34 countries or regions; n = 477 | UPCR at 9 months (interim); total eGFR slope over 24 months (final) | Total slope over 24 months | Mixed model for repeated measures / random-coefficients slope | Discontinuation of the study drug handled with a treatment-policy strategy; values after new background IgA nephropathy therapy excluded; missing values assumed missing at random | NEJM 2026 final report (Table 2) |
| NefIgArd (targeted-release budesonide | Phase 3, double-blind, placebo-controlled; 20 countries; n = | Time-weighted average eGFR | Total slope over 2 years (prespecified supportive | Random-coefficients model (supplementary | Full analysis set; missing data imputed by multiple imputation; sensitivity analyses with varied | Lancet 2023 appendix Table S3 (slope 1.82); |

| Trial (agent) | Design and sites | Primary end point | Slope end point and window | Slope model | Intercurrent events and missing data | Source of the slope estimate used |
|---|---|---|---|---|---|---|
| 16 mg daily for 9 months) | 364 | over 2 years | analysis) | Table S3) | missing-data assumptions | 2-year change/2 (2.95) used in sensitivity analysis |
| ALIGN (atrasentan 0.75 mg daily) | Phase 3, double-blind, placebo-controlled; 133 sites in 20 countries; main stratum n = 340 (SGLT2 inhibitor stratum n = 64 analysed separately) | UPCR at week 36 | Total slope over 136 weeks (key secondary); chronic slope (two-piece model, knot week 6) also reported | Random slope model with baseline UPCR, treatment, region, time and treatment-by-time | Data after intercurrent events (restricted or alternative IgA nephropathy medication, SGLT2 inhibitor initiation in the main stratum) censored; ITT supplementary analysis censoring only after SGLT2 inhibitor initiation consistent | Lancet 2026 appendix Table S1 (main stratum) |
| PROTECT (sparsentan 400 mg daily vs irbesartan 300 mg daily; sensitivity analysis only) | Phase 3, double-blind, double-dummy, active-controlled; 134 sites in 18 countries; n = 404 | UPCR at week 36 | Total slope day 1 to week 110; chronic slope week 6 to 110 | Mixed-effects slope model | Full analysis set; missing data imputed by multiple imputation under missing at random; modified ITT sensitivity analysis consistent | Lancet 2023 (Table 2) |

ITT, intention to treat; SC, subcutaneous; UPCR, urine protein-to-creatinine ratio.

#### Supplemental Table 2. Sensitivity analyses of the difference in total eGFR slope versus placebo

Values are mean differences in ml/min per 1.73 m2 per year (95% CI) with P-scores; common-effect network meta-analysis.

| Analysis | Telitacicept | Atacicept | Sibeprenlimab | Iptacopan | Targeted-release budesonide | Atrasentan | Sparsentan | Narsoplimab |
|---|---|---|---|---|---|---|---|---|
| Main analysis (six placebo-controlled trials) | 5.1 (3.5 to 6.7); 0.86 | 5.0 (3.6 to 6.5); 0.85 | 4.7 (3.8 to 5.6); 0.78 | 3.0 (2.0 to 4.0); 0.49 | 1.8 (0.5 to 3.1); 0.30 | 1.4 (0.5 to 2.3); 0.22 | - | - |
| Exclusion of conference data (TELIGAN) | - | 5.0 (3.6 to 6.5); 0.92 | 4.7 (3.8 to 5.6); 0.87 | 3.0 (2.0 to 4.0); 0.59 | 1.8 (0.5 to 3.1); 0.35 | 1.4 (0.5 to 2.3); 0.26 | - | - |
| Addition of PROTECT (irbesartan treated as placebo) | 5.1 (3.5 to 6.7); 0.88 | 5.0 (3.6 to 6.5); 0.87 | 4.7 (3.8 to 5.6); 0.81 | 3.0 (2.0 to 4.0); 0.56 | 1.8 (0.5 to 3.1); 0.37 | 1.4 (0.5 to 2.3); 0.29 | 1.0 (0.0 to 1.9); 0.20 | - |
| Slope windows of about 2 years only (exclusion of ALIGN) | 5.1 (3.5 to 6.7); 0.84 | 5.0 (3.6 to 6.5); 0.82 | 4.7 (3.8 to 5.6); 0.74 | 3.0 (2.0 to 4.0); 0.39 | 1.8 (0.5 to 3.1); 0.22 | - | - | - |
| Exclusion of NefIgArd | 5.1 (3.5 to 6.7); 0.84 | 5.0 (3.6 to 6.5); 0.82 | 4.7 (3.8 to 5.6); 0.74 | 3.0 (2.0 to 4.0); 0.41 | - | 1.4 (0.5 to 2.3); 0.20 | - | - |
| Chronic slope where reported (VISIONARY, ALIGN) | 5.1 (3.5 to 6.7); 0.89 | 5.0 (3.6 to 6.5); 0.87 | 4.3 (3.3 to 5.3); 0.73 | 3.0 (2.0 to 4.0); 0.49 | 1.8 (0.5 to 3.1); 0.27 | 1.7 (0.7 to 2.8); 0.25 | - | - |

| Analysis | Telitacicept | Atacicept | Sibeprenlimab | Iptacopan | Targeted-release budesonide | Atrasentan | Sparsentan | Narsoplimab |
|---|---|---|---|---|---|---|---|---|
| NefIgArd slope derived from the 2-year change (2.95) | 5.1 (3.5 to 6.7); 0.86 | 5.0 (3.6 to 6.5); 0.84 | 4.7 (3.8 to 5.6); 0.78 | 3.0 (2.0 to 4.0); 0.43 | 3.0 (1.5 to 4.4); 0.42 | 1.4 (0.5 to 2.3); 0.17 | - | - |
| ORIGIN 3 post hoc intention-to-treat slope (4.15) | 5.1 (3.5 to 6.7); 0.91 | 4.2 (2.8 to 5.5); 0.72 | 4.7 (3.8 to 5.6); 0.85 | 3.0 (2.0 to 4.0); 0.50 | 1.8 (0.5 to 3.1); 0.30 | 1.4 (0.5 to 2.3); 0.22 | - | - |
| Addition of ARTEMIS-IGAN (registry results, unverified) | 5.1 (3.5 to 6.7); 0.88 | 5.0 (3.6 to 6.5); 0.87 | 4.7 (3.8 to 5.6); 0.81 | 3.0 (2.0 to 4.0); 0.56 | 1.8 (0.5 to 3.1); 0.35 | 1.4 (0.5 to 2.3); 0.27 | - | 1.2 (-0.7 to 3.1); 0.24 |

### Supplemental Table 3. Risk of bias (RoB 2) for the total eGFR slope

| Trial | D1 Randomization | D2 Deviations from intended interventions | D3 Missing outcome data | D4 Measurement of the outcome | D5 Selection of the reported result | Overall | Main reasons |
|---|---|---|---|---|---|---|---|
| TELIGAN | Low | Low | High | Low | Some concerns | High | eGFR values after rescue or prohibited therapy were set to missing; such therapy was started in 33 of 159 placebo and 2 of 159 telitacicept participants by week 39, is triggered by rising proteinuria and falling eGFR, and no sensitivity analysis for missing eGFR data was reported (D3). Results available only as a conference poster, so agreement with the final statistical analysis plan could not be verified (D5). To be re-assessed on full publication. |
| ORIGIN 3 | Low | Low | Low | Low | Low | Low | Prespecified key secondary end point with treatment-policy strategy; full reporting with sensitivity analyses |
| VISIONARY | Low | Low | Low | Low | Low | Low | Prespecified hybrid estimand; treatment-policy sensitivity analysis consistent |
| APPLAUSE-IgAN | Low | Low | Low | Low | Low | Low | Prespecified key secondary end point; intercurrent-event handling prespecified and reported |
| NefIgArd | Low | Low | Low | Low | Low | Low | The slope estimate used is a prespecified supportive analysis in the appendix; the choice among reported estimates was made by the review according to a prespecified rule |
| ALIGN | Low | Low | Some concerns | Low | Low | Some concerns | eGFR censored after intercurrent events in |

| Trial | D1 Randomization | D2 Deviations from intended interventions | D3 Missing outcome data | D4 Measurement of the outcome | D5 Selection of the reported result | Overall | Main reasons |
|---|---|---|---|---|---|---|---|
| | | | | | | | 24% of the placebo group and 16% of the atrasentan group, mainly initiation of an SGLT2 inhibitor, with no sensitivity analysis reported for the total slope; the uncensored analysis of the change in eGFR at week 136 gave a smaller difference (D3). Prespecified key secondary end point through week 136. |
| PROTECT (sensitivity analysis) | Low | Low | Low | Low | Low | Low | Double-blind, double-dummy; prespecified slope analyses with multiple imputation |

Assessed with RoB 2 for the effect of assignment to intervention on the total eGFR slope by two reviewers; disagreements were resolved by discussion. The signalling-question answers and supporting quotations are available from the corresponding author. All trials were double-blind with central randomization and laboratory-measured eGFR.

#### Supplemental Table 4. Definitions of the composite kidney outcomes and proteinuria remission end points

| Trial | Composite kidney outcome (as reported) | Events, agent vs control | HR (95% CI) | Proteinuria remission end point (as reported) | Agent vs control |
|---|---|---|---|---|---|
| TELIGAN | Confirmed 30% or greater decline in eGFR through week 104 (40% or greater: HR 0.19, 0.06 to 0.56) | 4 vs 26 | 0.11 (0.04 to 0.30) | Not reported on the poster | - |
| ORIGIN 3 | Sustained 30% or greater decline in eGFR, sustained eGFR below 15 ml/min per 1.73 m2, long-term dialysis, kidney transplantation or death | 11 vs 38 | 0.24 (0.12 to 0.48) | Not reported | - |
| VISIONARY | 40% decline in eGFR or kidney failure composite through 24 months | 1 vs 28 | 0.03 (0.004 to 0.21) | Urinary protein excretion below 0.5 g/day (below 0.3 g/day) at 24 months | 36.7% vs 9.1% (22.9% vs 5.0%) |
| APPLAUSE-IgAN | Sustained 30% or greater decline in eGFR or kidney failure composite through 24 months | Hazard ratio reported | 0.57 (0.40 to 0.81) | 24-h UPCR below 1 g/g at 24 months | 40.7% vs 23.7% |
| NefIgArd | Confirmed 30% reduction in eGFR or kidney failure through 2 years | 21 vs 39 | 0.45 (0.26 to 0.75) | Not reported | - |
| ALIGN | 30% or greater decline in eGFR composite through week 136 | 26 vs 37 | 0.64 (0.39 to 1.05) | Partial remission (below 1 g/day) reported as an end point; values not extracted | - |
| PROTECT (vs irbesartan) | Confirmed 40% reduction in eGFR, end-stage kidney disease or all-cause mortality through week 110 | 18 vs 26 | Relative risk 0.7 (0.4 to 1.2) | Complete remission below 0.3 g/day; below 0.5 g/day (post hoc); partial remission below | 31% vs 11% (RR 2.5, 1.6 to 4.1); 51% vs 24% (RR 2.1, |

| Trial | Composite kidney outcome (as reported) | Events, agent vs control | HR (95% CI) | Proteinuria remission end point (as reported) | Agent vs control |
|---|---|---|---|---|---|
| | | | | 1.0 g/day | 1.5 to 2.9); 78% vs 53% (RR 1.5, 1.1 to 1.9) |

Because the definitions differ between trials, the hazard ratios were pooled only to examine the direction and consistency of the effects (Supplemental Figure 7), and the proteinuria remission end points were not pooled.

#### Supplemental Table 5. Within-trial subgroup estimates of the difference in total eGFR slope versus placebo

Values are ml/min per 1.73 m2 per year (95% CI) from the subgroup analyses reported by each trial (ORIGIN 3: supplementary Figure S6 of the final report; VISIONARY: Figure 2 of the final report, slope from week 4; APPLAUSE-IgAN: Figure 2 of the final report; ALIGN: appendix of the final report, main stratum). Proteinuria cut-off 1.5 g/g (VISIONARY 2.0 g/g); ALIGN eGFR 45 or higher combined from the 45-60 and above-60 strata; VISIONARY regions combined by inverse variance (Asia = East Asia and South or Southeast Asia).

| Subgroup | Level | ORIGIN 3 (atacicept) | VISIONARY (sibeprenlimab) | APPLAUSE-IgAN (iptacopan) | ALIGN (atrasentan) |
|---|---|---|---|---|---|
| Proteinuria | High UPCR | 6.3 (4.2 to 8.3) | 5.9 (4.0 to 7.9) | 2.3 (1.1 to 3.6) | 2.4 (1.0 to 3.9) |
| Proteinuria | Low UPCR | 3.9 (1.9 to 5.8) | 4.2 (3.2 to 5.3) | 3.2 (1.8 to 4.7) | 0.8 (-0.4 to 2.0) |
| eGFR | eGFR below 45 | 4.6 (2.5 to 6.6) | 3.7 (2.3 to 5.2) | 2.4 (0.7 to 4.0) | 1.5 (-0.0 to 3.1) |
| eGFR | eGFR 45 or higher | 5.2 (3.3 to 7.1) | 4.7 (3.6 to 5.8) | 3.3 (2.0 to 4.5) | 1.4 (0.3 to 2.6) |
| Region | Asia | 5.0 (2.9 to 7.2) | 5.4 (4.0 to 6.7) | 2.5 (1.1 to 3.9) | 1.6 (0.2 to 3.0) |
| Region | Other regions | 5.0 (3.0 to 6.9) | 3.3 (2.2 to 4.4) | 3.5 (2.1 to 4.9) | 1.2 (-0.0 to 2.4) |
| Sex | Male | 5.6 (3.6 to 7.6) | 4.3 (3.1 to 5.5) | 2.9 (1.5 to 4.3) | 1.3 (0.1 to 2.6) |
| Sex | Female | 4.0 (2.0 to 6.0) | 4.9 (3.5 to 6.2) | 3.3 (2.0 to 4.7) | 1.7 (0.3 to 3.0) |
| Age (cut-off 40, 40, 45 years) | Younger | 6.6 (4.2 to 9.0) | 5.6 (4.1 to 7.1) | 2.7 (1.4 to 4.0) | - |
| Age (cut-off 40, 40, 45 years) | Older | 3.7 (2.0 to 5.4) | 3.7 (2.6 to 4.8) | 3.7 (2.2 to 5.2) | - |
| SGLT2 inhibitor at baseline | Yes | 5.1 (3.4 to 6.8) | 4.8 (3.5 to 6.1) | 4.0 (1.8 to 6.1) | 3.0 (0.9 to 5.0) |
| SGLT2 inhibitor at baseline | No | 4.8 (2.2 to 7.4) | 4.3 (3.0 to 5.6) | 2.7 (1.6 to 3.8) | 1.4 (0.5 to 2.3) |

Pooled within-trial differences (inverse variance, common effect): proteinuria, high minus low, 1.0 (-0.1 to 2.0), I2 = 44%, P = 0.07; eGFR, below 45 minus 45 or higher, -0.6 (-1.6 to 0.4), I2 = 0%, P = 0.26; region, Asia minus other, 0.6 (-0.4 to 1.6), I2 = 45%, P = 0.26; sex, male minus female, -0.2 (-1.2 to 0.8), I2 = 0%, P = 0.74; age, younger minus older, 0.9 (-0.3 to 2.2), I2 = 70%, P = 0.13 (B-cell-directed agents only, 2.2, 0.6 to 3.8); SGLT2 inhibitor, receiving minus not receiving, 0.9 (-0.2 to 2.1), I2 = 0%. ALIGN SGLT2 inhibitor values are the SGLT2 inhibitor stratum versus the main stratum.

#### Supplemental Table 6. League table of the main analysis

Difference in total eGFR slope (ml/min per 1.73 m2 per year, 95% CI), row treatment versus column treatment; positive values favour the row treatment. Comparisons between active agents are indirect.

| | Telitacicept | Atacicept | Sibeprenlimab | Iptacopan | TRF-budesonide | Atrasentan | Placebo |
|---|---|---|---|---|---|---|---|
| Telitacicept | **Telitacicept** | | | | | | 5.1 (3.5 to 6.7) |
| Atacicept | 0.1 (-2.1 to 2.3) | **Atacicept** | | | | | 5.0 (3.6 to 6.5) |
| Sibeprenlimab | 0.4 (-1.4 to 2.2) | 0.3 (-1.4 to 2.0) | **Sibeprenlimab** | | | | 4.7 (3.8 to 5.6) |
| Iptacopan | 2.1 (0.2 to 4.0) | 2.0 (0.2 to 3.7) | 1.7 (0.3 to 3.0) | **Iptacopan** | | | 3.0 (2.0 to 4.0) |
| TRF-budesonide | 3.3 (1.2 to 5.4) | 3.2 (1.2 to 5.1) | 2.9 (1.3 to 4.5) | 1.2 (-0.4 to 2.8) | **TRF-budesonide** | | 1.8 (0.5 to 3.1) |
| Atrasentan | 3.7 (1.9 to 5.5) | 3.6 (1.9 to 5.3) | 3.3 (2.0 to 4.6) | 1.6 (0.3 to 3.0) | 0.4 (-1.2 to 2.0) | **Atrasentan** | 1.4 (0.5 to 2.3) |
| Placebo | 5.1 (3.5 to 6.7) | 5.0 (3.6 to 6.5) | 4.7 (3.8 to 5.6) | 3.0 (2.0 to 4.0) | 1.8 (0.5 to 3.1) | 1.4 (0.5 to 2.3) | **Placebo** |

TRF-budesonide, targeted-release budesonide. Values in the lower triangle are column treatment minus row treatment as reported by netmeta (row placebo: active agent versus placebo).

#### Supplemental Table 7. Certainty of evidence (CINeMA) for the difference in total eGFR slope

Difference in total eGFR slope in ml/min per 1.73 m2 per year (95% CI) from the main network meta-analysis; positive values favour the first-named treatment. Comparisons with placebo are direct (one trial each); comparisons between active agents are indirect.

| Comparison | Difference (95% CI) | Within-study bias | Reporting bias | Indirectness | Imprecision | Heterogeneity | Incoherence | Confidence |
|---|---|---|---|---|---|---|---|---|
| Telitacicept vs placebo | 5.1 (3.5 to 6.7) | Major concerns | Low risk | No concerns | No concerns | No concerns | No concerns | Low |
| Atacicept vs placebo | 5.0 (3.6 to 6.5) | No concerns | Low risk | No concerns | No concerns | No concerns | No concerns | High |
| Sibeprenlimab vs placebo | 4.7 (3.8 to 5.6) | No concerns | Low risk | No concerns | No concerns | No concerns | No concerns | High |
| Iptacopan vs placebo | 3.0 (2.0 to 4.0) | No concerns | Low risk | No concerns | No concerns | No concerns | No concerns | High |
| TRF-budesonide vs placebo | 1.8 (0.5 to 3.1) | No concerns | Low risk | No concerns | No concerns | No concerns | No concerns | High |
| Atrasentan vs placebo | 1.4 (0.5 to 2.3) | Some concerns | Low risk | No concerns | No concerns | No concerns | No concerns | Moderate |
| Telitacicept vs atacicept | 0.1 (-2.1 to 2.3) | Some concerns | Low risk | Some concerns | Major concerns | No concerns | No concerns | Very low |
| Telitacicept vs sibeprenlimab | 0.4 (-1.4 to 2.2) | Some concerns | Low risk | Some concerns | Major concerns | No concerns | No concerns | Very low |
| Atacicept vs sibeprenlimab | 0.3 (-1.4 to 2.0) | No concerns | Low risk | Some concerns | Major concerns | No concerns | No concerns | Very low |
| Telitacicept vs iptacopan | 2.1 (0.2 to 4.0) | Some concerns | Low risk | Some concerns | No concerns | No concerns | No concerns | Low |
| Atacicept vs iptacopan | 2.0 (0.2 to 3.7) | No concerns | Low risk | Some concerns | No concerns | No concerns | No concerns | Moderate |
| Sibeprenlimab vs iptacopan | 1.7 (0.3 to 3.0) | No concerns | Low risk | Some concerns | No concerns | No concerns | No concerns | Moderate |
| Telitacicept vs TRF-budesonide | 3.3 (1.2 to 5.4) | Some concerns | Low risk | Some concerns | No concerns | No concerns | No concerns | Low |
| Atacicept vs TRF-budesonide | 3.2 (1.2 to 5.1) | No concerns | Low risk | Some concerns | No concerns | No concerns | No concerns | Moderate |
| Sibeprenlimab vs TRF-budesonide | 2.9 (1.3 to 4.5) | No concerns | Low risk | Some concerns | No concerns | No concerns | No concerns | Moderate |
| Iptacopan vs TRF-budesonide | 1.2 (-0.4 to 2.8) | No concerns | Low risk | Some concerns | Some concerns | No concerns | No concerns | Low |
| Telitacicept vs atrasentan | 3.7 (1.9 to 5.5) | Major concerns | Low risk | Some concerns | No concerns | No concerns | No concerns | Very low |
| Atacicept vs atrasentan | 3.6 (1.9 to 5.3) | Some concerns | Low risk | Some concerns | No concerns | No concerns | No concerns | Low |
| Sibeprenlimab vs atrasentan | 3.3 (2.0 to 4.6) | Some concerns | Low risk | Some concerns | No concerns | No concerns | No concerns | Low |
| Iptacopan vs atrasentan | 1.6 (0.3 to 3.0) | Some concerns | Low risk | Some concerns | No concerns | No concerns | No concerns | Low |
| TRF-budesonide vs atrasentan | 0.4 (-1.2 to 2.0) | Some concerns | Low risk | Some concerns | Major concerns | No concerns | No concerns | Very low |

CI, confidence interval; TRF-budesonide, targeted-release budesonide. Ratings were made with the CINeMA web application (version 3.0.1; fixed-effect model, mean difference). Within-study bias: the consensus RoB 2 judgement of each trial (Supplemental Table 3) summarized across the contribution matrix by the average rule; major concerns for the direct comparison informed by TELIGAN (high risk of bias) and some concerns for comparisons informed by ALIGN (some concerns) or partly by TELIGAN. Reporting bias: low risk for all comparisons because trial registries and conference abstracts were searched and slope results were available for every included trial. Indirectness: no concerns at the study level; comparisons between active agents were rated as some concerns because they rest on the assumption of transitivity across trials that differed in region, background SGLT2 inhibitor use, length of the slope window, and slope methodology (Table 1 and Supplemental Table 1). Imprecision, heterogeneity, and incoherence were judged with a clinically important difference of 0.75 ml/min per 1.73 m2 per year (set by the review team and informed by the treatment effects on GFR slope associated with clinical benefit in the meta-analysis by Inker et al., Nat Med 2023): imprecision was rated as some concerns when the 95% CI included no effect and extended into clinically important effects in one direction, and as major concerns when it extended into clinically important effects in both directions. The estimated between-study variance was 0, so prediction and confidence intervals coincided, and incoherence could not be tested because each comparison was informed by a single trial and the network had no closed loops; both domains were rated as no concerns. Confidence started at high and was rated down by one level for each domain with some concerns and two levels for each domain with major concerns. SGLT2, sodium-glucose cotransporter 2.

#### Supplemental Figures

**Supplemental Figure 1.** PRISMA 2020 flow diagram of study selection (databases, registers and other sources).

**Supplemental Figure 2.** Network of the main analysis: six placebo-controlled trials, each contributing one comparison with placebo (star-shaped network); node size proportional to the number of randomized participants.

**Supplemental Figure 3.** Risk of bias (RoB 2) for the total eGFR slope in each trial.

**Supplemental Figure 4.** Difference in total eGFR slope versus placebo by baseline proteinuria (a), baseline eGFR (b), region (c), and sex (d) in ORIGIN 3, VISIONARY, APPLAUSE-IgAN, and ALIGN, with the pooled within-trial difference between subgroups.

**Supplemental Figure 5.** Difference in total eGFR slope versus placebo by age (younger versus older, trial-specific cut-off of 40 or 45 years) in ORIGIN 3, VISIONARY, and APPLAUSE-IgAN.

**Supplemental Figure 6.** Difference in the change in eGFR from baseline at the end of the slope window versus placebo (network meta-analysis of four trials).

**Supplemental Figure 7.** Hazard ratios versus placebo for the composite kidney outcome of each trial (definitions in Supplemental Table 4). KF, kidney failure; KRT, kidney replacement therapy.

### Supplemental Figure 1

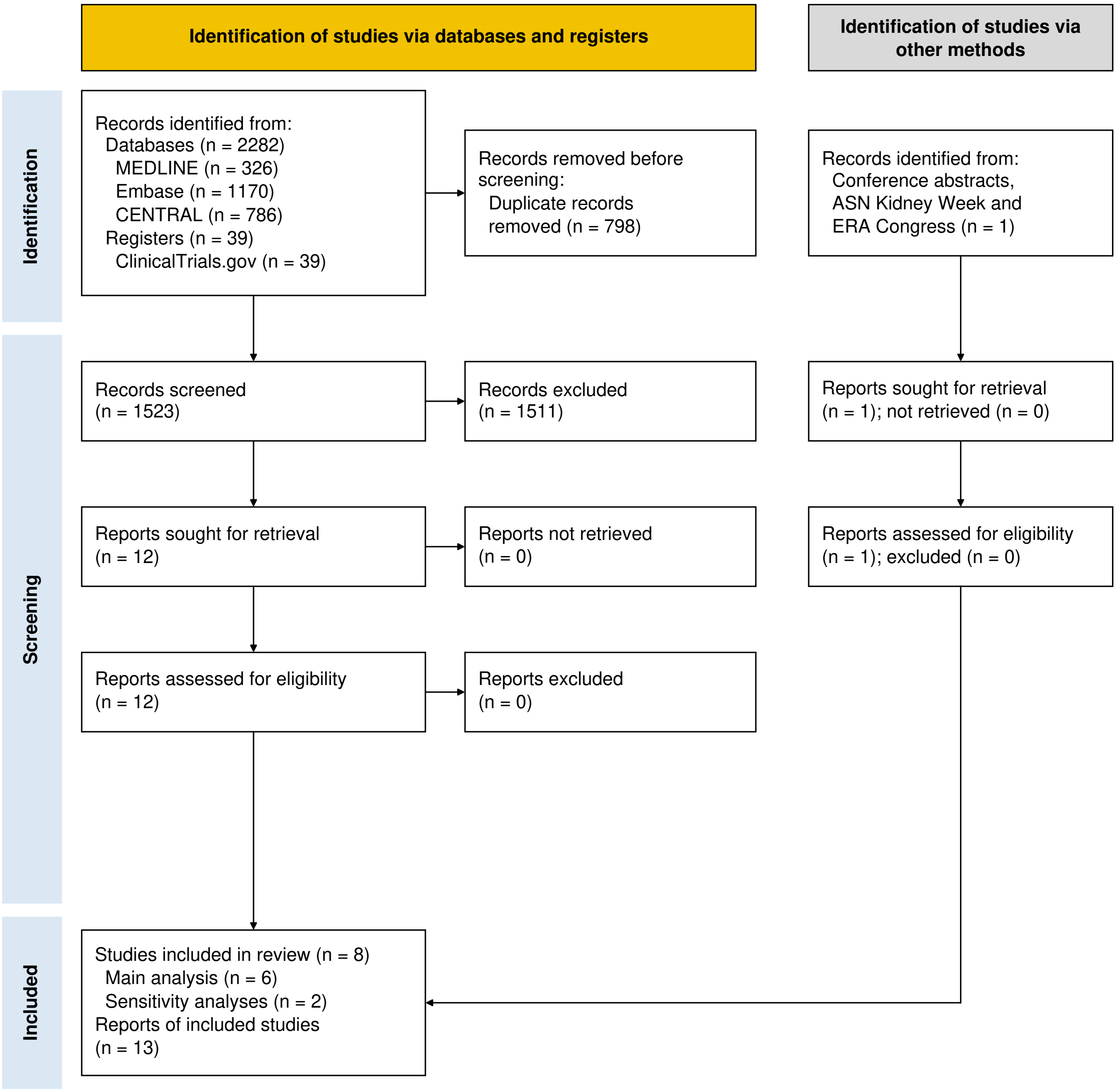

### Supplemental Figure 2

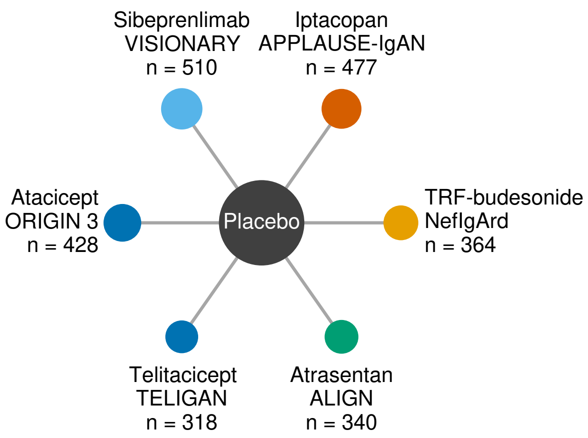

### Supplemental Figure 3

| | D1 | D2 | D3 | D4 | D5 | Overall |
|---|---|---|---|---|---|---|
| Telitacicept (TELIGAN) | + | + | X | + | - | X |
| Atacicept (ORIGIN 3) | + | + | + | + | + | + |
| Sibeprenlimab (VISIONARY) | + | + | + | + | + | + |
| Iptacopan (APPLAUSE-IgAN) | + | + | + | + | + | + |
| TRF-budesonide (NefIgArd) | + | + | + | + | + | + |
| Atrasentan (ALIGN) | + | + | - | + | + | - |
| Sparsentan (PROTECT, sensitivity) | + | + | + | + | + | + |

D1 Randomization process
D2 Deviations from intended interventions
D3 Missing outcome data
D4 Measurement of the outcome
D5 Selection of the reported result

\+ Low - Some concerns x High

### Supplemental Figure 4

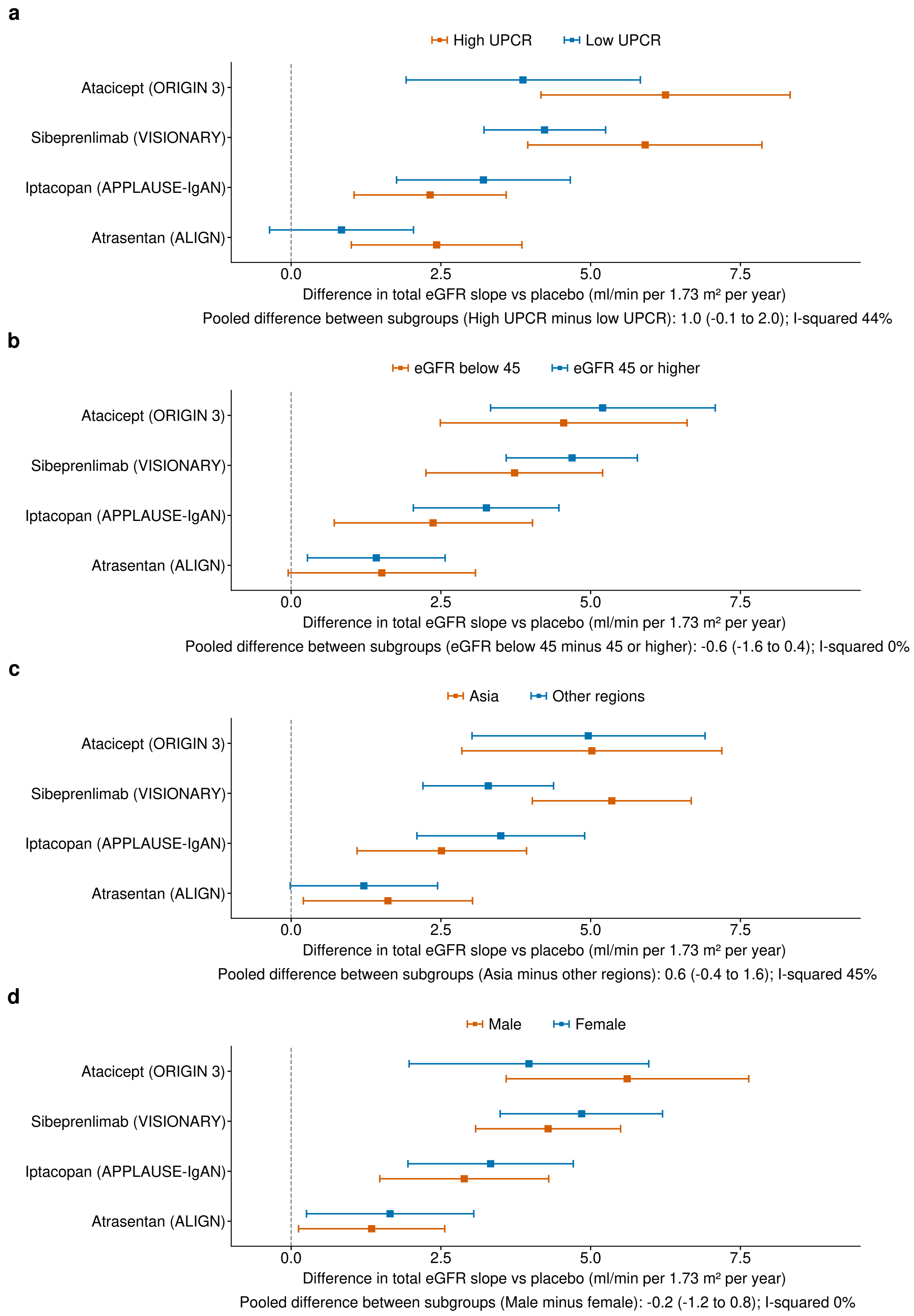

### Supplemental Figure 5

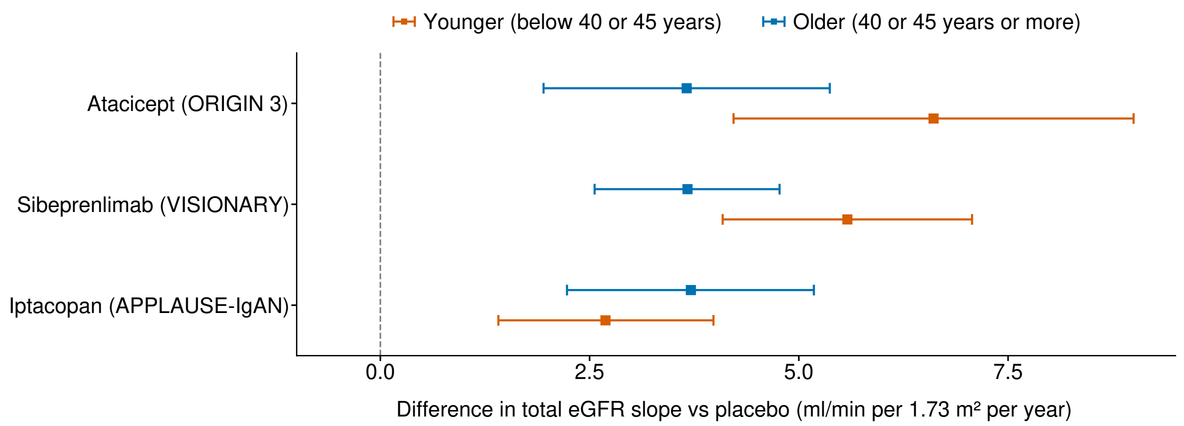


Pooled difference (younger minus older): all three trials 0.9 (-0.3 to 2.2), I-squared 70%; B-cell-directed agents only 2.2 (0.6 to 3.8)

### Supplemental Figure 6

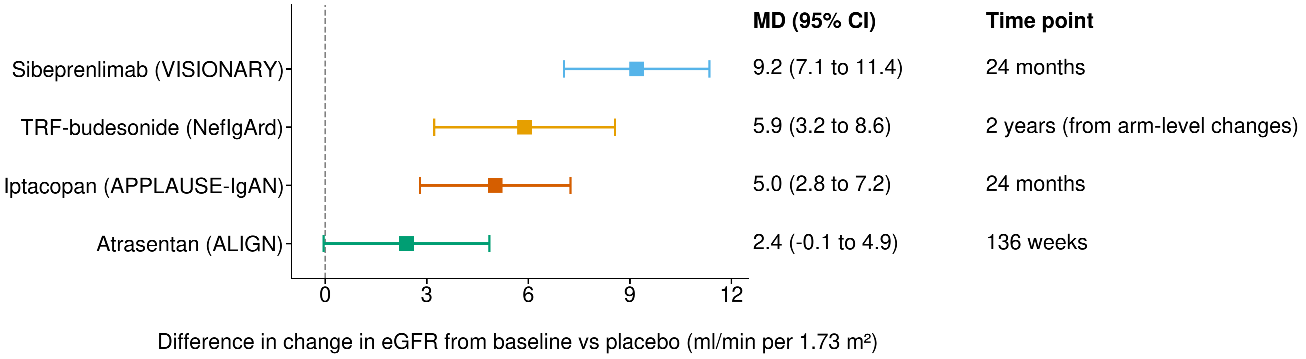

### Supplemental Figure 7

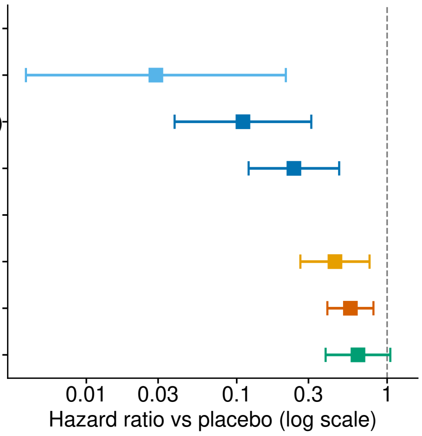


| | HR (95% CI) | Outcome in the trial |
|---|---|---|
| **B-cell-directed agents** | | |
| Sibeprenlimab (VISIONARY) | 0.03 (0.004 to 0.21) | 40% decline or KF |
| Telitacicept (TELIGAN) | 0.11 (0.04 to 0.31) | 30% decline (confirmed) |
| Atacicept (ORIGIN 3) | 0.24 (0.12 to 0.48) | 30% decline, eGFR <15, KRT or death |
| **Other disease-targeted agents** | | |
| TRF-budesonide (NefIgArd) | 0.45 (0.26 to 0.76) | 30% decline (confirmed) or KF |
| Iptacopan (APPLAUSE-IgAN) | 0.57 (0.40 to 0.81) | 30% decline (sustained) or KF |
| Atrasentan (ALIGN) | 0.64 (0.39 to 1.05) | 30% decline composite |
